# Factors influencing wellbeing in young people during COVID-19: A survey with 6291 young people in Wales

**DOI:** 10.1101/2021.08.13.21261959

**Authors:** Michaela James, Hope Jones, Amana Baig, Emily Marchant, Tegan Waites, Charlotte Todd, Karen Hughes, Sinead Brophy

## Abstract

COVID-19 infection and the resultant restrictions has impacted all aspects of life across the world. This study explores factors that promote or support wellbeing for young people during the pandemic, how they differ by age, using a self-reported online survey with those aged 8 - 25 in Wales between September 2020 and February 2021. Open-ended responses were analysed via thematic analysis to provide further context. A total of 6,291 responses were obtained from 81 education settings across Wales (including primary and secondary schools as well as sixth form, colleges and universities). Wellbeing was highest in primary school children and boys and lowest in those who were at secondary school children, who were girls and, those who preferred not to give a gender. Among primary school children, higher wellbeing was seen for those who played with others (rather than alone), were of Asian ethnicity (OR 2.3, 95% CI: 1.26 to 4.3), lived in a safe area (OR: 2.0, 95% CI: 1.67 to 2.5) and had more sleep. To support their wellbeing young people reported they would like to be able to play with their friends more. Among secondary school children those who were of mixed ethnicity reported lower wellbeing (OR: 5.10, 95% CI: 1.70 to 15.80). To support their wellbeing they reported they would like more support with mental health (due to anxiety and pressure to achieve when learning online). This study found self-reported wellbeing differed by gender, ethnicity and deprivation and found younger children report the need for play and to see friends to support wellbeing but older children/young people wanted more support with anxiety and educational pressures.

## Introduction

As a result of the COVID-19 global pandemic declared by the World Health Organization in March 2020 (1), a wide range of guidance and restrictions on movement were implemented by governments worldwide in order to reduce transmission. In the United Kingdom (UK) the government enforced a national ‘lockdown’ with a strict ‘stay at home’ message consisting of the closure of education settings, non-essential retail and leisure and formal working spaces. This also included the restriction of social gatherings and the use of outdoor space to exercise (2,3).

The economic cost of the pandemic, school closures and lockdown for the government has been considerable. However, there are a number of other potential costs that need to be considered namely that of population wellbeing (4). The instructions to stay at home have impacted the physical and mental health of the population (5). Many will have experienced fears and concerns such as the risk of infection, loss of social contacts, and confinement (6).

Despite being deemed the population least vulnerable to direct harms from COVID infection (7), the government enforced restrictions impacted the lives of young people in an unprecedented way (8). In particular, education closures saw a disruption to daily routines, reduced opportunities for physical activity, lack of socialisation and an adaptation to remote learning, which all could have detrimental impacts on physical and mental health and wellbeing (9–12). Feelings of isolation, stress, anxiety and unhappiness have been noted during the pandemic (8) as support networks (friends, education settings, sports clubs and mental health support) were unable to operate under restrictions (13). As a result, the home environment played a pivotal role in young people’s lives becoming the place for support, socialisation, education and activity as well as isolating children from external support (14).

The pandemic may have long-term consequences on young people (8). Recent reports from children’s charities have suggested how wide and varying these may be as perceptions of the pandemic range from a period of self-reflection and re-connecting with families to feeling isolated and concerned over their futures (13). These varying experiences have been reflected in Welsh data (15). However, a more recent report by Young Minds suggests that as the pandemic continues, the detrimental impacts on young people’s mental health is becoming more significant with time (16).

It is unclear what the full significance of these consequences are. However, there is a growing body of evidence examining young people’s health and wellbeing during the pandemic (17–19). The aim of this study is to explore predictors of wellbeing for children and young people during COVID, providing recommendations that span from primary school to higher education. Learning from this can help identify groups who are most at risk of low wellbeing and need further support. This is particularly relevant as lockdown is eased and services begin to resume. This identification and support could be invaluable for mitigating the long-term consequences on children and young people, as well as identifying recommendations that can be utilised given any future lockdowns.

## Materials and Methods

### Study design

This study utilised data collected by the Healthy Development team in the National Centre for Population Health and Wellbeing Research (NCPHWR) between September 2020 and February 2021. Two online surveys facilitated data collection; the ‘HAPPEN’ Survey and the ‘COVID & Young People’ Survey (supplementary file 1).

The primary aim of this study was to identify the predictors of wellbeing for children and young people during the COVID 19 pandemic. As a secondary aim, qualitative data from open ended responses in both questionnaires were explored giving children and young people a voice to express what influenced their wellbeing during the pandemic. Therefore, this study provides key predictors and recommendations to improve wellbeing for this age group.

### Participants

For the HAPPEN Survey, recruitment of participants (aged 8 – 11) was done online due to COVID-19 restrictions. Pre-existing HAPPEN schools within the network were emailed initially inviting them to participate in the survey. The survey was then opened wider and all primary schools in Wales were contacted through several methods including direct email and promotion from key stakeholders (e.g., regional education consortia). The survey questions can be seen as supplementary file 1.

A similar approach was taken for the COVID & Young People survey (aged 11 – 25) by directly emailing education settings and stakeholders to promote the survey to participants. In addition, a social media campaign (paid advertisement on Facebook and Twitter) was also used for wider recruitment.

### Data collection

Data was collected between September 2020 and February 2021 lasting for 5 months. This encompassed a period of education setting closure between 14^th^ December 2020 to the end of February 2021. The HAPPEN Survey is an annual survey for primary school children aged 8–11 years delivered through the HAPPEN Wales network; a network which brings together education, health and research in line with the new curriculum proposals for health and wellbeing Wales. HAPPEN is novel as it operates a data collection and feedback system which is achieved by sharing group-level results to schools as a school report tailored to the curriculum (20). The HAPPEN survey is an online self-report questionnaire that was developed and designed with children. The survey captures a range of information on health and wellbeing including nutrition, physical activity, sleep, wellbeing and concentration. Prior to COVID-19, children completed the survey within the school setting during curriculum time. However, due to school closures some data collection was carried out in the home setting. The survey was granted ethical approval by Swansea University’s Medical School on 15/04/2020 (Reference: 2017-0033B).

The COVID & Young People survey recruited those aged from 11 – 25 from secondary, sixth form/college and university settings. Like the HAPPEN survey, it captured a range of health and wellbeing information and was designed alongside young people and stakeholders. Participants completed the survey in their own time, independently. The COVID & Young People study was granted ethical approval by Swansea University Medical School on the 14^th^ October 2020 (Ref: 2020-0049).

Both surveys captured the typical health behaviours of children and young people aged 8-25. Items included validated measures of health and wellbeing. For this study, the Me and My Feelings (MMF) measure of emotional and behavioural difficulties was used for those under 11 through the HAPPEN Survey and the Generalised Anxiety Disorder (GAD7) measure used for those aged 11-25 (secondary school, sixth form/college, university) through the COVID & Young People Survey. The Good Childhood Index (developed by the Children’s Society) was included in both surveys and measured subjective wellbeing. Participants were asked to rank their happiness for a number of indicators including their health, life, education, family and friends from 1 – 10 (10 being the most happy). There were some differences in the questions asked for example, the HAPPEN Survey explicitly asked about play (supplementary file 1).

The MMF measure covers two domains: emotional and behavioural difficulties. For this study, both domains were used to describe wellbeing. The MMF measure determines these categories by assigning scores of 0, 1 and 2 to responses of ‘Never’, ‘Sometimes’ and ‘Always’ to 16 items. These items are summed to give an overall score. Cut-offs established by Deighton et al. (2013), define the Emotional Difficulties Subscale (questions 1 – 11). Scores of 10 and 11 indicate borderline difficulties, and scores of 12 and above indicate clinically significant difficulties. For the Behavioural Difficulties Subscale (questions 11 – 16). Scores of 6 indicate borderline difficulties, and scores of 7 and above indicate clinically significant difficulties. Emotional and behavioural difficulties in this study represent a classification of clinical.

The GAD7 assigns scores of 0, 1, 2, and 3, to the categories of ‘*not at all*’, ‘*several days*’, *‘more than half the days*’, and ‘*nearly every day*’ and sums the scores for the 7 questions. Scores of 5, 10, and 15 are taken as the cut-off points for mild, moderate and severe anxiety, respectively. For both measures, the continuous and binary scales were considered for analysis. For the binary outcome, a score of 1 was assigned to those displaying clinical scores and 0 assigned to anything below.

The process of data coding involved 2 researchers. The first researcher (MJ) downloaded the raw data from both surveys, cleaned the data and generated a unique participant ID number before removing identifiable information. This process protects participants’ anonymity. Raw data was coded using STATA (version 16) to produce a dataset for the purpose of analyses. Open-ended survey questions were further coded by a second researcher (HJ).

### Analysis

Quantitative analysis was undertaken by 3 researchers in April 2021. Initially, the lead researcher (MJ) examined the predictors of wellbeing by education setting. Logistic regression was used to analyse binary data (adjusting for age). Linear regression was used to explore any statistically significant differences between education settings and measures on the Good Childhood Index. Factors associated with emotional difficulties in primary school aged children and for general anxiety in teenagers/young people were analyzed using backwards stepwise logistic regression. All factors were included in the initial model and those with a significance value of >0.1 were removed manually one-by-one until the final model contained only factors with a significance of <0.1.

A qualitative thematic analysis (TA) approach was used to explore common themes from the open-ended responses to the HAPPEN survey and COVID & Young People survey. This was undertaken by a researcher (HJ) to explore common themes emerging from the responses. Codebook thematic analysis was used to generate themes from an open-ended questions from the surveys. For the HAPPEN Survey, this included “*If you could change something to make you and your friends healthier and happier, what would you change*… *IN SCHOOL/OUT OF SCHOOL*” and in the COVID & Young People Survey this included the “*Why is this?*” (supplementary file 1). Braun and Clarke’s Phases of TA (2006) (21) underpinned the coding process. Each response was read and assigned an initial code. Each code was then categorised and themes identified from these codes in an inductive process. From these themes, sub-themes were then explored. This process was reviewed by a second researcher on the project for validation (MJ). The researchers used TA to identify and report patterns in the focus groups.

## Results

The combined surveys had a total of 6,291 responses from 81 education settings across Wales including primary and secondary schools as well as sixth form, colleges and universities. The flow of responses can be seen as figure 1.

**Figure 1-.**
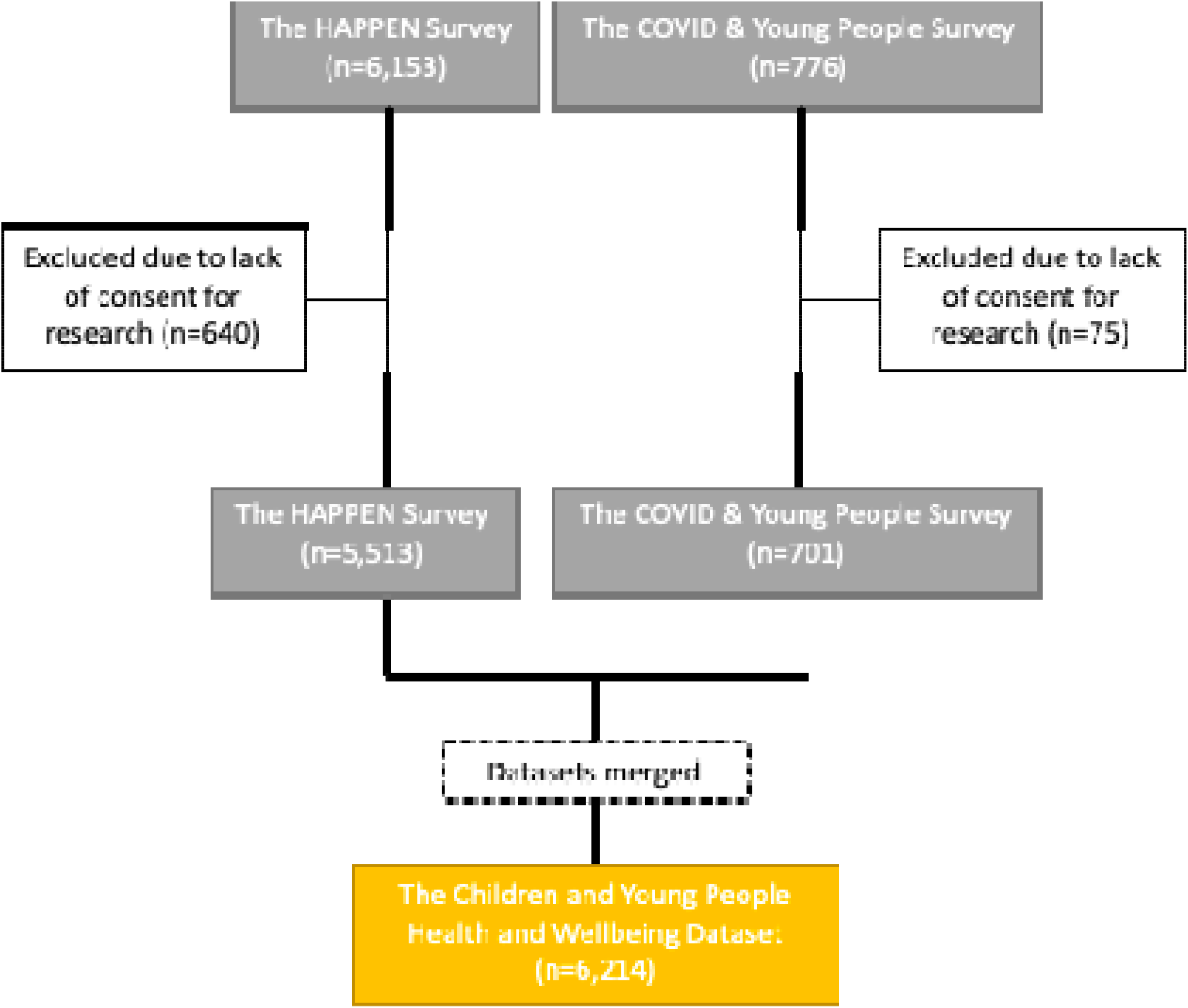
Flow of participant responses.

**Figure 2-.**
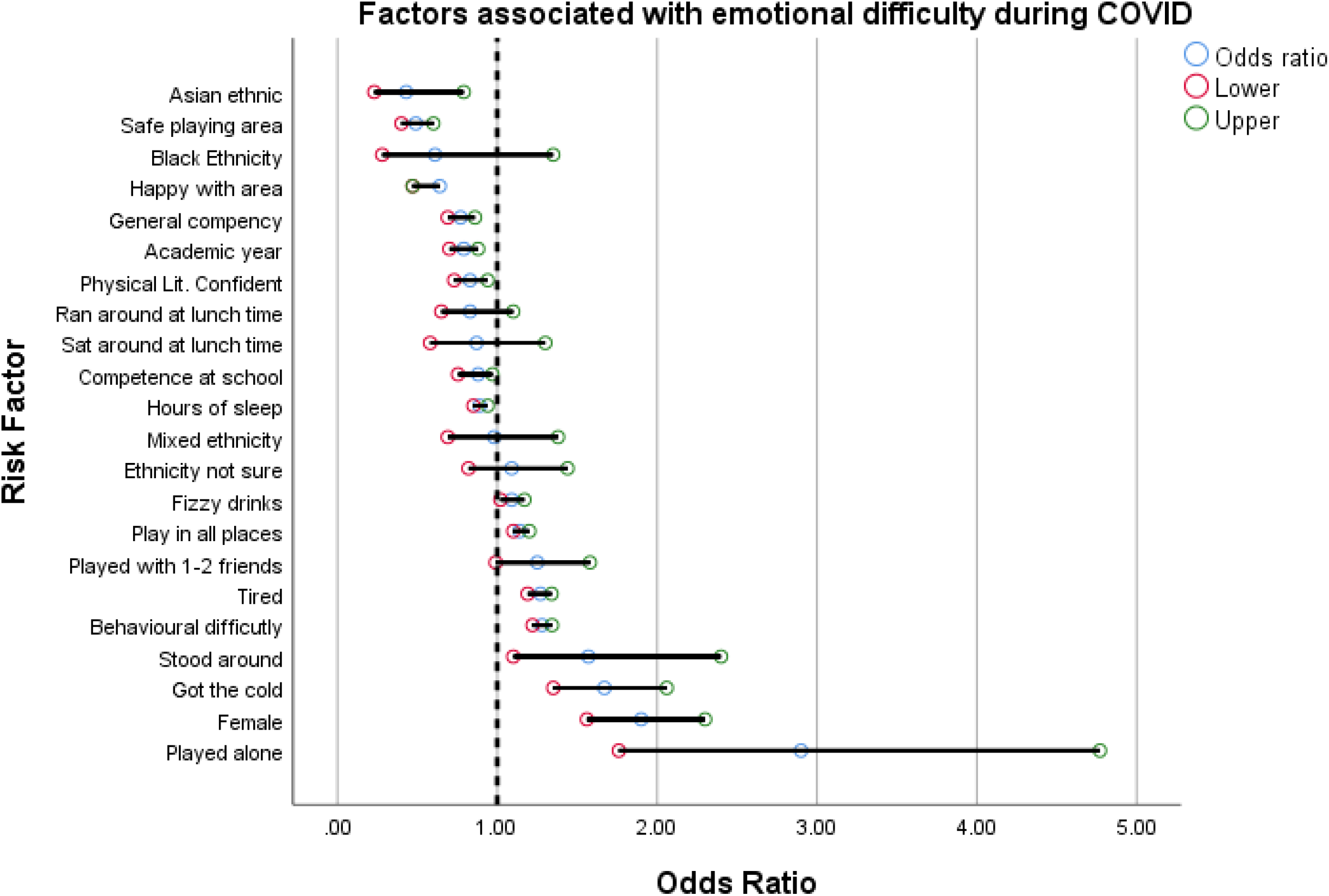
Factors associated with emotional difficulties in children during COVID.

**Fig. 3.**
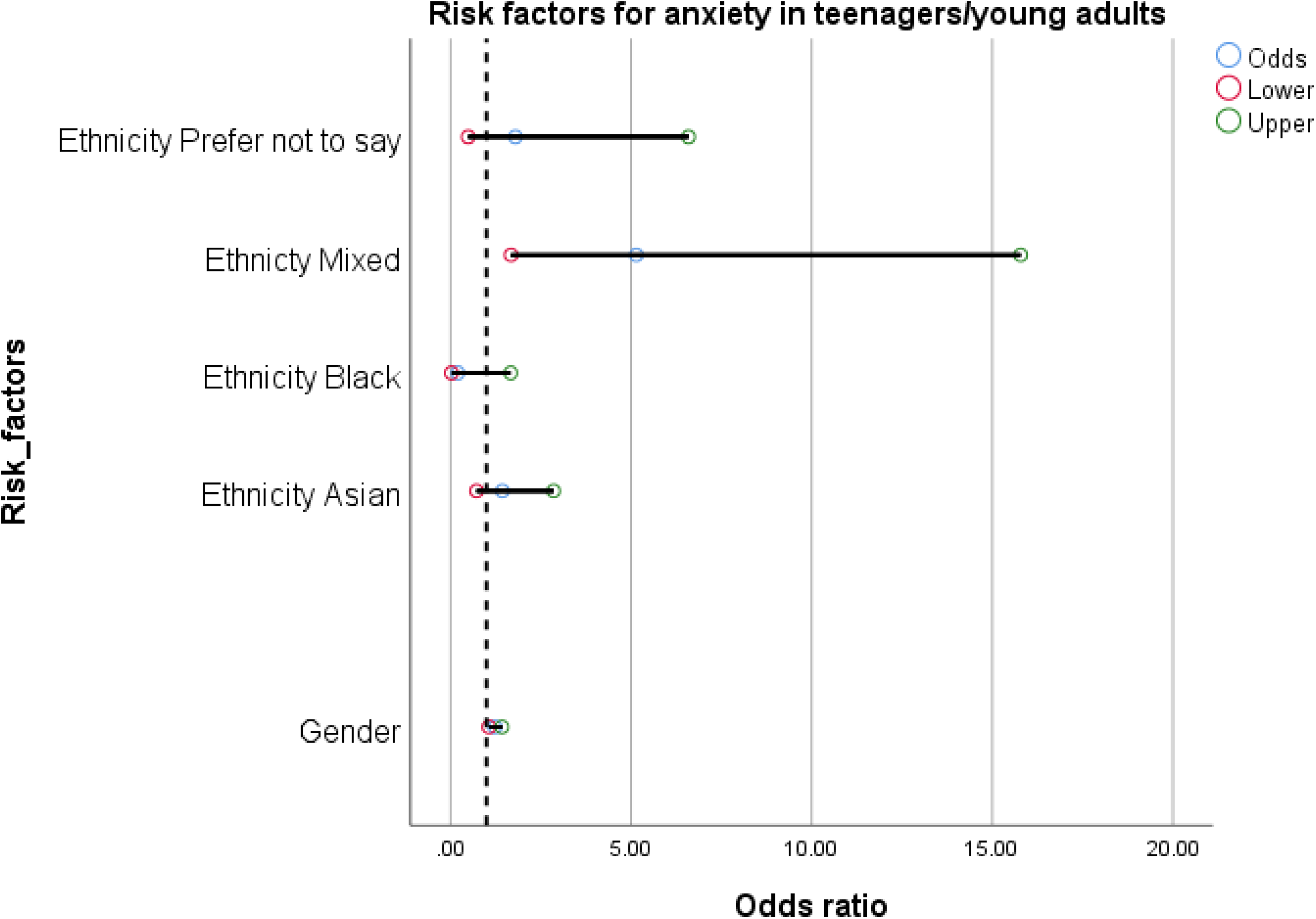
Factors associated with anxiety in teenagers/young adults.

*Fig. 1 – Flow of responses*

A breakdown of demographics of study participants by education setting can be seen as table 1.

**Table 1-.** Demographics

|  | Group (n=6214) | Primary (n=5513) | Secondary (n=401) | Sixth Form/College (n=197) | University (n=103) |
| --- | --- | --- | --- | --- | --- |
|  | Freq. (%) / Mean (SD) | Freq. (%) / Mean (SD) | Freq. (%) / Mean (SD) | Freq. (%) / Mean (SD) | Freq. (%) / Mean (SD) |
| Age | 10.7 (2.39) | 10.04 (.8) | 13.90 (1.40) | 17.5 (.7) | 21.4 (3.8) |
| Gender |  |  |  |  |  |
| Girl | 3,159 (50.21) | 2,969 (48.90%) | 208 (51.87) | 139 (70.59) | 69 (66.99) |
| Boy | 2,725 (43.31) | 2,488 (45.13%) | 143 (35.66) | 48 (24.37) | 23 (22.33) |
| Prefer not to say | 269 (4.28) | 244 (4.43%) | 16 (3.99) | 5 (2.52) | 1 (0.97) |
| Missing | 139 (2.21) | 85 (1.54%) | 34 (8.48) | 5 (2.52) | 10 (9.71) |
| Ethnicity |  |  |  |  |  |
| Asian | 232 (3.69%) | 187 (3.39%) | 22 (5.49) | 11 (5.58) | 9 (8.74) |
| Black | 115 (1.83%) | 99 (1.79%) | 2 (0.50) | 2 (1.02) | 9 (8.74) |
| Mixed | 422 (6.72%) | 399 (7.25%) | 9 (2.25) | 11 (5.59) | 6 (5.82) |
| White | 4,695 (74.49%) | 4,039 (73.39%) | 347 (86.53) | 168 (85.28) | 74 (71.84) |
| Prefer not to say | 712 (11.34%) | 695 (12.62%) | 14 (3.49) | 1 (0.51) | 1 (0.97) |
| Missing | 101 (1.60%) | 94 (1.70%) | 7 (1.25) | 4 (2.03) | 4 (3.88) |
| Deprivation (WIMD Quintile(22)) |  |  |  |  |  |
| 1 (most deprived) | 25.35% | 26.61% | 14.89% | 18.35% | 10.14% |
| 2 | 14.84% | 14.08% | 22.70% | 18.99% | 15.94% |
| 3 | 15.93% | 15.23% | 24.47% | 20.25% | 17.39% |
| 4 | 23.90% | 23.94% | 21.99% | 21.52% | 30.43% |
| 5 (least deprived) | 19.99% | 20.14% | 15.96% | 20.89% | 26.09% |
| Wellbeing (Me and My Feelings) |  |  |  |  |  |
| Emotional Difficulties |  | 6.78 (4.00) |  |  |  |
| Behavioural Difficulties |  | 3.01 (2.48) |  |  |  |
| Anxiety (GAD7) |  |  |  |  |  |
| Anxiety Score | 7.20 (6.44) |  | 7.18 (5.99) | 10.68 (5.79) | 9.35 (6.09) |
| Life Measures (1-10) (Good Childhood Index) |  |  |  |  |  |
| Physical Health | 7.59 (2.64) | 8.08 (2.23) | 3.99 (2.70) | 4.03 (2.81) | 4.6 (2.73) |
| Education | 8.32 (2.33) | 8.43 (2.26) | 7.09 (2.80) | 8.54 (2.06) | 7.53 (2.52) |
| Family | 8.92 (2.08) | 9.29 (1.68) | 6.11 (2.75) | 6.57 (2.47) | 6.36 (2.87) |
| Friends | 8.56 (2.20) | 8.89 (1.91) | 5.63 (2.72) | 6.67 (2.45) | 6.91 (2.46) |
| Life | 8.36 (2.35) | 8.53 (2.28) | 6.94 (2.54) | 7.64 (2.31) | 7.8 (2.03) |

The demographic table (table 1) shows that participation rates were much higher at primary school, where there was an even gender split. The rates decrease as participants age and the gender split weighs in favour of girls. Level of deprivation reduces through education setting with primary settings reporting higher deprivation than university settings. As well as this, fewer participants of black ethnicity were participating from higher educational settings. Mean scores for self-reported happiness with health, education, family, friends, and life all decrease between primary and secondary school participants.

Of those of primary school age 11.96% presented clinical emotional difficulty scores. These scores were significantly higher in girls with 13.53% (compared to 8.67% in boys, 95% CI: - 6.64 to -3.07). In older children and young people, 21.21% of participants reported high scores anxiety scores. Again, this was much higher in girls (25.73% compared to 8.55%, 95% CI: -24.03 to -10.32).

### Factors associated with self-reported happiness across education settings

Table 2 shows the difference in scores as ranked by participants using the Good Childhood Index. Differences are presented using an age-graded comparison reference group of the previous education stream (e.g., secondary refers to comparison of secondary with primary).

**Table 2–.** Differences in self-reported happiness compared to primary aged children

|  |  |  |  |
| --- | --- | --- | --- |
| Physical Health |  | Difference | 95% CI |
|  | Secondary | -4.08 | **-4.33 to -3.85** |
|  | College/Sixth Form | .04 | -.35 to .44 |
|  | University | .56 | -.00 to 1.12 |
| Education |  |  |  |
|  | Secondary | -1.33 | **-1.57 to -1.08** |
|  | College/Sixth Form | 1.44 | 1.04 to 1.84 |
|  | University | .43 | -.08 to .94 |
| Family |  |  |  |
|  | Secondary | -3.17 | **-3.37 to -2.98** |
|  | College/Sixth Form | .46 | .14 to .78 |
|  | University | .24 | -.16 to .65 |
| Friends |  |  |  |
|  | Secondary | -3.26 | **-3.48 to -3.05** |
|  | College/Sixth Form | 1.04 | **.69 to 1.39** |
|  | University | 1.28 | **.82 to 1.73** |
| Life |  |  |  |
|  | Secondary | -1.58 | **-1.82 to -1.34** |
|  | College/Sixth Form | .70 | **.30 to 1.10** |
|  | University | .85 | **.33 to 1.37** |
| *bold denotes significant difference between means |  |  |  |

Significant differences were seen in all indicators of self-reported happiness between primary and secondary school, suggesting a decline in happiness between these two age groups. Happiness with health (−4.08, CI: -4.33 to -3.85) and happiness with friends (−3.26, CI: -3.48 to -3.05) show the biggest declines. This then increased from secondary school to further education (get older) suggesting those in higher education have higher happiness across indicators. Therefore, table 2 suggests that being in secondary school during the pandemic is a predictor of poorer wellbeing (self-reported happiness). However, this may also be the case in normal circumstances too.

### Specific predictors associated with emotional difficulties at aged 8-11 (primary school) using the HAPPEN Survey

Logistic regression models showed children who self-reported emotional difficulties during COVID, were more likely to be those who reported playing alone more (OR: 2.9, 95% CI: 1.76 to 4.77, compared to those who play with 4 or more others), girls (OR: 1.9, 95% CI: 1.56 to 2.30, compared to boys), those who had a cold or COVID symptoms (OR: 1.67, 95% CI: 1.35 to 2.06) and feel tired (OR: 1.27, 95% CI: 1.19 to 1.34), those who had behavioral difficulties according to MMF (OR: 1.29, 95% CI: 1.22 to 1.34, for each 1 point increase in overall score), those who reported standing around at break time when in school (OR 1.57, 95% CI: 1.10 to 2.4, compared to those who walked around) and those who reported drinking more fizzy drinks over 7 days (OR: 1.09, 95% CI: 1.02 to 1.17). Factors associated with lower risk of emotional difficulties included; Asian ethnicity (OR 0.43, 95% CI: 0.23 to 0.79 compared to white ethnicity), perceiving their local area (near their home) as safe to play (OR: 0.49, 95% CI: 0.40 to 0.60), feeling happy in the area they live (OR: 0.64, 95% CI: 0.47 to 0.47), being in an older academic year (OR: 0.79, 95% CI: 0.70 to 0.88), feeling confident in terms of physical literacy (OR: 0.83, 95% CI: 0.75 to 0.94) and having more sleep (OR: 0.89, 95% CI: 0.85 to 0.94) for each hour of extra sleep). See Table 3.

**Table 3–.** Specific factors associated with emotional difficulty in children aged 9 - 11

|  |  |  |
| --- | --- | --- |
| Risk Factor | Odds | 95% CI |
| Played Alone | 2.90 | **1.76 to 4.77** |
| Female | 1.90 | **1.56 to 2.30** |
| Got A Cold/COVID-19 Symptoms | 1.67 | **1.35 to 2.06** |
| Stood Around At Breaktime | 1.57 | **1.1 to 2.4** |
| Behavioural Difficulty Score | 1.28 | **1.22 to 1.342** |
| Tired | 1.27 | **1.19 to 1.34** |
| Played With 1-2 Friends | 1.25 | *0.99 to 1.58* |
| Play In All Places | 1.14 | **1.1 to 1.2** |
| Fizzy Drinks | 1.09 | **1.02 to 1.17** |
| Ethnicity Not Sure<sup>s</sup> | 1.09 | 0.82 to 1.44 |
| Mixed Ethnicity<sup>s</sup> | 0.98 | 0.69 to 1.38 |
| Hours Of Sleep | 0.89 | **0.85 to 0.94** |
| Competence At School | 0.88 | **0.754 to 0.97** |
| Sat Around At Lunchtime | 0.87 | 0.58 to 1.3 |
| Ran Around Lunchtime | 0.83 | 0.65 to 1.1 |
| Physical Literacy Confident | 0.83 | **0.73 to 0.94** |
| Academic Year | 0.79 | **0.7 to 0.88** |
| General Competence | 0.77 | **0.69 to 0.86** |
| Happy With Area | 0.64 | **0.47 to 0.47** |
| Black Ethnicity<sup>§</sup> | 0.61 | 0.28 to 1.35 |
| Safe Playing Area | 0.49 | **0.4 to 0.6** |
| Asian Ethnicity<sup>§</sup> | 0.43 | **0.23 to 0.79** |
| <sup>§</sup> Compared to white ethnicity. |  |  |

Factors which were considered but were removed from the model due to lack of significant association with emotional difficulty were; deprivation of the area, ability to ride a bike or swim, ability to walk to the park, how the child travels to or from school, diet (breakfast, fruit and veg, sugary snacks, number of takeaways), number of times brushing teeth, physical activity of 60 minutes a day, number of hours sedentary, afternoon break in school, having a garden.

### Specific factors associated with anxiety in teenagers/young adults (secondary to university)

Young people with higher general anxiety score were more likely to be those who were mixed ethnicity (average GAD score for those of mixed ethnicity: 13.6 compared to 8.6 for those of white ethnicity and 10.5 for those who reported ‘prefer not to say’) (table 1) and girls (average GAD score for girls was 9.8 compared to 5.8 for boys and 11.5 for those who reported ‘prefer not to say’). See table 4.

**Table 4–.** Specific factors associated with anxiety in teenagers/young adults

|  |  |  |
| --- | --- | --- |
| Risk Factor | Odds Ratio | 95% CI |
| Female | **1.20** | **1.10 to 1.40** |
| Asian<sup>§</sup> | 1.43 | 0.70 to 2.80 |
| Black<sup>§</sup> | 0.20 | 0.02 to 1.66 |
| Mixed<sup>§</sup> | 5.10 | **1.70 to 15.80** |
| Prefer Not To Say | 1.80 | 0.50 to 6.60 |
| <sup>§</sup>Compared to white ethnicity |  |  |

Factors investigated but removed from the model due to lack of significant association with anxiety included; number of adults in the household, wearing a mask (associated with a trend of higher anxiety score) and ability to social distance.

### Qualitative results: Self-reported influences of wellbeing for children and young people during the pandemic

A total of 6292 responses were analysed from the open-ended responses; 5513 responses were from primary schools and 777 from secondary school to university. Three main themes emerged from primary school responses; physical health, being with friends and coronavirus concerns. In secondary school to university, responses equated to 3 main themes with subsequent sub-themes; mental health support (the negative impact of COVID-19), exam pressure and uncertainty (assessment concerns, feeling behind and need for support, learning from home, lack of motivation and positives of distance learning) and future prospects. These themes can be seen as supplementary file 3.

Primary school pupils highlight that to make themselves and their friends happier they want to improve their physical health by doing more physical activity. They mention that they would like; “*Longer time to play and exercise*”, “*Healthy eating*” and to “*Play in the park*”. This was relevant in the school setting where they highlighted they wanted “*More PE*” suggesting they wanted more opportunities to be active across the school day. As well as this, they asked for “*More sports equipment*”. It is unclear whether this refers to many different types of equipment or simply more of one item. Some children also cited improving their physical health by “*Healthy eating*” or “*Eat more fruit and veg*” acknowledging this would also make them happier.

As well as this, primary aged children wanted to be able to socialise with their friends more as a way of making them happier. For example, they say they want to “*Live closer to my friends*”, “*Be with my friends more*” and “*Being able to play places I want to with my friends*”. The latter quote shows how being physically active in the form of play and being with friends are linked for younger children and these opportunities to play with others is important for their happiness.

This age group showed concerns around coronavirus saying that they wanted “*For the virus to go away*” and to “*Stop the spread of covid-19 so we can get back to normal*” including for the restrictions in place to be ended; “*No more covid rules*”. Coupled with the responses above this is likely to be because of the restrictions around physical activity and socialisation making children less happy throughout the pandemic.

Due to the wording of the open-ended questions in the surveys, older children and young people’s responses focused around exam pressure and the uncertainty coronavirus restrictions have caused. In particularly, they noted there needs to be an increase in mental health support namely that the forward focus of education settings should be on wellbeing and mental health over educational attainment in the return to education after the pandemic. For example, one participant stated; “*School for me was never easy but now more than ever that pressure to achieve without the understanding that myself and peers are struggling through an incredibly difficult time and we need to prioritise ourselves and self-care before we even consider the notion of exams in such daunting times*”. They describe heightened anxiety and stress during the pandemic with comments like; “*The worsened depression and heightened anxiety caused by coronavirus has almost pushed me to drop out of sixth form*”.

However, the main stressors behind this need for additional support were around assessment concerns and feeling behind particularly for secondary school pupils. In particular, “*Exam cancellation, predicted grades changed, stress of university choices without having to visit them. Unsure of my future in education*”. They reported that they felt they had been left in the dark about the exam situation which keeps changing. For example, “*Don’t know what is happening with exams etc*.”, “*Exam situation is very stressful for everyone at the moment, lack of clarity and answers which is currently needed as we have missed so much time off school and deserve to know what will happen and how it might affect our future*.” This has been exacerbated by working from home and working online. This centered around comments such as, “*Things are much more difficult when you are trying to learn from home*”. In particular, staying motivated was a major concern which has an impact on schoolwork as they described the differences between in-person and online teaching; “*Online work is not nearly as effective as in person learning*” and “*I struggle immensely with learning remotely. It’s too easy to get distracted by other things*”. They also stress the exam pressure they are under and feeling behind in work; “*Pressure to well in GCSE’s despite missing a lot of school time*.”

Some university students highlight that the pandemic makes it harder to prepare for life after university including applying for further study or looking for a job. In particular, “*It has made me less motivated to properly prepare for leaving university i*.*e. sorting out a job, postgraduate studies etc*.” and “*My chances of securing a graduate job have fallen dramatically and could potentially force me into another career, wasting my degree*.”

However, it is important to note that some individuals at university reported a positive impact of education during coronavirus having enjoyed learning from home and working online. In particular; “*I actually find online learning better, because I can replay the recorded lesson back and make notes in my own time*”, “*Online learning is a good alternative to face to face and I appreciate that it is a useful tool*” and “*My university is truly doing it’s very best to provide the same level of education as always*.”

Similarly, to secondary school students, they reported difficulty with online learning. Notably in terms of the lack of support; “*I feel like I am doing some online course with almost no support or a sense of community. Feels like I am all alone trying to get my degree*.”. In parallel to the younger children, university students also note missing socialisation and being with others. For example, “*Lack of social interactions with course mates and other friends*”. They reported a much wider impact of the changes due to COVID; “*You are trapped, you can’t do what you usually do. This has affected my education, my physical and mental health*”.

The noticeable change is that older children become more aware of their future and long-term goals; older children displaying concerns for their future, their education, exams and future prospects after the pandemic. In comparison, younger children live predominantly in the present and want to be with their friends and play.

## Discussion

When examining children and young people as a group, it is evident that being a girl and being in secondary school is a predictor of reporting feeling less happy (with health, education, life, family, and friends), having clinical emotional difficulties and higher anxiety. Research shows that girls are more likely to experience internalising difficulties than boys and this gender gap increases as young people age (23). In a wider context, studies focusing on mental health during COVID-19 in particular note that women and young people have experienced worse mental health outcomes (24). However, it has also been noted that wellbeing deteriorated amongst young people as a whole during this time period (25).

With our study showing poorer wellbeing among girls in secondary school, greater consideration is needed over appropriate avenues of support for these target grounds. It is evident from wider research that both gender (as well as socioeconomic inequalities) have been widened during the pandemic (25). This needs to be noted and work needs to be done to seek to close this gap or stop it widening further. Social and emotional learning programs have shown good evidence of effectiveness in reducing symptoms of depression and anxiety in the short-term and could be used as potential interventions (26). A recent report by Children in Wales suggests young people felt there has been increased waiting lists, increased time to get support and less communication about how to get mental health-based support during the pandemic (27). Therefore, the provision of services during the return to school and adequate signposting to available services could provide opportunities to regain support they may have lost or not had access to.

Specific predictors of emotional difficulties in primary school children suggest that playing alone, reporting having a cold (e.g., not feeling well), being less active at break time, consuming more fizzy drinks and feeling tired result in higher odds of these difficulties. Results of this study suggests that emotional wellbeing could be improved for primary aged children if they are supported in socialising and feeling confident and competent. The main factor that they reported affecting their wellbeing in the qualitative analysis was the ability to play with friends and indeed playing alone was associated with emotional difficulties in the quantitative analysis. Whilst causality is difficult to determine due to the nature of this study, giving opportunities to play and supporting play opportunities in COVID recovery plans could help improve emotional wellbeing.

This was complimented by the qualitative analysis which highlighted the need for play and socialisation in younger children. Ensuring children have a safe area to play will be important to the wellbeing of primary school children during and post COVID. The benefits of play and being physically active include cognitive, physical, social and emotional development (28). During the pandemic, the benefits of play have been impacted due to prolonged periods where the ability to play for all ages has been compromised (29) as access to resources, facilities and friendship groups have been restricted. This means the benefits of this activity could have been mitigated. This was mirrored in school staff recommendations made in a previous study which noted that teachers observed pupils’ weight gain, lethargy, anxiety, low mood and social disconnection (30) upon the return to school as they felt children were engaging in less physical activity. Wider research has suggested that lockdown restrictions saw children (aged 3 – 12) unable to leave their homes and concluded that not only did this have a considerable impact on wellbeing at a psychological level but also in social and physical terms with children reporting missing their peers and opportunities to be active outside (17). A recent report highlighted that behavioural and emotional difficulties in children, as well as anxiety and stress in caregivers has increased throughout the pandemic with current findings surpassing that of the first lockdown (31); perhaps as a result of the negative links between parental stress and child wellbeing (32). This suggests that attention needs to be paid to the longitudinal effect of rolling lockdowns as ‘stress points’ for family support systems.

Previous research by HAPPEN on self-reported health behaviour measures from the first lockdown (April – June 2020) from children (aged 8 – 11) showed that children, as a group, were actually more physically active (33). Whilst this provides a contrasting narrative, those on free school meals (used as a proxy for deprivation) who reported being less physically active. This highlights an inequality in provision/access which may have been widened during lockdown. Findings from the current study emphasis the importance of protecting play, socialisation and opportunities to be active (particularly in younger children) is paramount to avoid widening any inequalities and ensuring that wellbeing remains a priority.

Findings from older children and young people suggest anxiety is associated with ethnicity (identifying as mixed ethnicity) and that those who prefer not to say their gender or ethnicity had higher anxiety levels. In a wider context, there is evidence emerging around ethnicity-specific changes in mental health during the pandemic (34) with a higher increase in mental distress seen in ethnic minority groups. It has also noted that this increase is gender specific; with ethnic minority men experiencing increases in mental distress yet women as a whole experience these increases in mental distress regardless of ethnicity during COVID-19 regardless of background (34). It is worth nothing that this previous research has been with an adult population, but this study does provide further evidence that there may be ethnicity-specific wellbeing influences during the pandemic. In contrast, research with primary-aged children shows that being of Asian, black of mixed ethnicity at primary school has been associated with less difficulties (35). A similar finding is present in this study with those being of Asian ethnicity having lower odds of a clinical emotional difficulty.

The impact on mental health and wellbeing was reflected in the qualitative findings which highlighted how older participants considered mental health support to be a significant recommendation during the return to pre-pandemic life. Age has been one of the clearest determinants of mental wellbeing with younger adults exhibiting poorer outcomes than older adults and younger peers (32,36). Work by Khan et al. (19) suggests this may be because of a lack of physical exercise, financial uncertainty, fear of infection, lack of information and guidance and excessive reporting of the pandemic on news outlets. In this study, teenagers expressed a need for more support with their mental health rather than educational attainment and the catch-up with learning.

It has been suggested that to reduce inequalities in education occurring in lockdown, children should engage with or seek ‘catch-up’/tuition particularly those from more deprived areas (37). Research shows that recommendations made by school staff highlight the need to remove the pressure placed on assessments/attainment and focus on wellbeing as a priority (30) particularly for younger pupils. It is evident that online learning was a cause of anxiety for many secondary school pupils. In particular, the lack of support from education during school closures. Secondary school is traditionally teacher-led in delivery style and as the move to more independent learning in lockdown, this may have proved a difficult transition for young people at a time of uncertainty; particularly for those in assessment years. In contrast, those at university expressed concerns about their prospects rather than their present circumstances. This study highlights the differences in wellbeing predictors throughout the ages.

However, despite the negative impact of COVID-19 on wellbeing particularly for girls and older children, qualitative results from this study suggest there may also have been some positives particularly with online learning. Some participants noted that they could learn better and at their own pace at home with less pressure to keep up with a class and they could have one to one support from a parent. Whilst it is important to take note the negative effects of the pandemic when learning for the future, the positives must also be taken forward.

### Limitations

Although the studies surveys were made available to all children (aged 8-11) and young people (aged 12 – 18+) in full-time/part-time education across Wales, the findings of this paper only present those who participated in the survey. Participation rates decreased throughout education settings as well as deprivation and participation from ethnic minority groups. Therefore, more research is needed to identify factors associated with wellbeing in children who have left school and left education.

Furthermore, due to the cross-sectional design of the study, direct comparisons to pre-pandemic wellbeing and emotional difficulties or anxiety were not made, and causal conclusions cannot be drawn. However, the study does provide some indication of which students may be most vulnerable to poorer wellbeing outcomes because of the pandemic or have been exacerbated because of the pandemic.

## Conclusion

When examining the predictors of wellbeing in children and young people as a group, it is evident that gender (being a girl) and age (being in secondary school) are significant predictors of feeling less happy (with health, education, life, family, and friends), having emotional difficulties and anxiety. This is not surprising given previous research, prior to the pandemic, which highlights these groups as at risk of poorer mental health outcomes. However, this study does provide evidence that needs to be noted by support services, schools and wider partners who can help close the gap within these inequalities.

More specific recommendations from this study highlight the need for play and socialisation in younger children and specific mental health and future support for those who are older. Thus, protecting play, socialisation, and opportunities to be active (particularly in younger children) is paramount to avoid widening any inequalities and ensuring that wellbeing remains a priority. It is evident that online learning was a cause of anxiety for secondary school pupils thus, the return to school needs to ensure a smooth transition back to face-to-face teaching with the need to remove the pressure placed on assessments/attainment and focus on wellbeing as a priority and support for prospects.

## Data Availability

No additional data available.

## Acknowledgements

The authors would like to thank the participants who took the time to engage with and complete our surveys during an unprecedented time and to thank the educational settings and organisations that also promoted the surveys.

## References

1. World Health Organisation (WHO). WHO announces COVID-19 outbreak a pandemic [Internet]. 2020 [cited 2021 Apr 29]. Available from: https://www.euro.who.int/en/health-topics/health-emergencies/coronavirus-covid-19/news/news/2020/3/who-announces-covid-19-outbreak-a-pandemic

2. Dawson DL, Golijani-Moghaddam N. COVID-19: Psychological flexibility, coping, mental health, and wellbeing in the UK during the pandemic. J Context Behav Sci [Internet]. 2020;17(July):126–34. Available from: https://doi.org/10.1016/j.jcbs.2020.07.010

3. British Broadcasting Company (BBC). Coronavirus: Strict new curbs on life in UK announced by PM [Internet]. [cited 2021 Apr 29]. Available from: https://www.bbc.co.uk/news/uk-52012432

4. Brodeur A, Clark AE, Fleche S, Powdthavee N. COVID-19, lockdowns and well-being: Evidence from Google Trends. J Public Econ [Internet]. 2021;193:104346. Available from: https://doi.org/10.1016/j.jpubeco.2020.104346

5. Thakur K, Kumar N, Sharma NR. Effect of the Pandemic and Lockdown on Mental Health of Children. Indian J Pediatr. 2020;87(7):552.

6. Wright LJ, Williams SE, Veldhuijzen van Zanten JJCS. Physical Activity Protects Against the Negative Impact of Coronavirus Fear on Adolescent Mental Health and Well-Being During the COVID-19 Pandemic. Front Psychol. 2021;12(March).

7. Clemens V, Deschamps P, Fegert JM, Anagnostopoulos D, Bailey S, Doyle M, et al. Potential effects of “social” distancing measures and school lockdown on child and adolescent mental health. Eur Child Adolesc Psychiatry [Internet]. 2020;29(6):739–42. Available from: https://doi.org/10.1007/s00787-020-01549-w

8. Singh S, Roy D, Sinha K, Parveen S, Sharma G, Joshi G. Impact of COVID-19 and lockdown on mental health of children and adolescents: A narrative review with recommendations. Psychiatry Res [Internet]. 2020;293(August):113429. Available from: https://doi.org/10.1016/j.psychres.2020.113429

9. Van Lancker W, Parolin Z. COVID-19, school closures, and child poverty: a social crisis in the making. Lancet Public Heal [Internet]. 2020;5(5):243–4. Available from: http://dx.doi.org/10.1016/S2468-2667(20)30084-0

10. Wang G, Zhang Y, Zhao J, Zhang J, Jiang F. Mitigate the effects of home confinement on children during the COVID-19 outbreak. Lancet. 2020;395(10228):945–7.

11. Liu JJ, Bao Y, Huang X, Shi J, Lu L. Mental health considerations for children quarantined because of COVID-19. Lancet Child Adolesc Heal. 2020;4(5):347–9.

12. Lyons RA, Jones KH, John G, Brooks CJ, Verplancke P, Ford D V, et al. BMC Medical Informatics and The SAIL databankl⍰: linking multiple health and social care datasets. 2009;8:1–8.

13. Day L, Percy-smith B, Rizzo S, Erskine C, Monchuk L, Shah M. To lockdown and back: Research report. 2020.

14. Amerio A, Brambilla A, Morganti A, Aguglia A, Bianchi D, Santi F, et al. Covid-19 lockdown: Housing built environment’s effects on mental health. Int J Environ Res Public Health. 2020;17(16):1–10.

15. Children’s Commissioner for Wales. Coronavirus and Me. 2020;1–68. Available from: https://www.mind.org.uk/information-support/legal-rights/coronavirus-and-your-rights/coronavirus-and-sectioning/

16. YoungMinds. Coronavirus: Impact on young people with mental health needs Survey 2: Summer 2020. YoungMinds [Internet]. 2020;(February):1–18. Available from: https://youngminds.org.uk/about-us/reports/coronavirus-impact-on-young-people-with-mental-health-needs/

17. Idoiaga Mondragon N, Berasategi Sancho N, Dosil Santamaria M, Eiguren Munitis A. Struggling to breathe: a qualitative study of children’s wellbeing during lockdown in Spain. Psychol Heal [Internet]. 2020;0(0):1–16. Available from: https://doi.org/10.1080/08870446.2020.1804570

18. Dodd RH, Dadaczynski K, Okan O, McCaffery KJ, Pickles K. Psychological wellbeing and academic experience of university students in australia during covid-19. Int J Environ Res Public Health. 2021;18(3):1–12.

19. Khan AH, Sultana MS, Hossain S, Hasan MT, Ahmed HU, Sikder MT. The impact of COVID-19 pandemic on mental health & wellbeing among home-quarantined Bangladeshi students: A cross-sectional pilot study. J Affect Disord [Internet]. 2020;277(August):121–8. Available from: https://doi.org/10.1016/j.jad.2020.07.135

20. Todd C, Chistian D, Tyler R, Stratton G, Brophy S. Developing HAPPEN (Health and Attainment of Pupils involved in a Primary Education Network): working in partnership to improve child health and education. Perspect Public Health. 2016;136(3):115–6.

21. Braun V, Clarke V. Using thematic analysis in psychology. Qual Res Psychol. 2006;3(2):77–101.

22. Statistics for Wales. Welsh Index of Multiple Deprivation. 2011;1–5. Available from: www.wales.gov.uk/statistics

23. Campbell OLK, Bann D, Patalay P. The gender gap in adolescent mental health: A cross-national investigation of 566,829 adolescents across 73 countries. SSM – Popul Heal [Internet]. 2021;13(January):100742. Available from: https://doi.org/10.1016/j.ssmph.2021.100742

24. Jacques-Avinõ C, López-Jiménez T, Medina-Perucha L, De Bont J, Goncąlves AQ, Duarte-Salles T, et al. Gender-based approach on the social impact and mental health in Spain during COVID-19 lockdown: A cross-sectional study. BMJ Open. 2020;10(11):1–10.

25. Ford T, John A, Gunnell D. Mental health of children and young people during pandemic. BMJ. 2021;372(March):1–2.

26. Clarke A, Sorgenfrei M, Mulcahy J, Davie P, Friedrich C, Mcbride T, et al. Adolescent mental health A systematic review on the effectiveness of school-based. 2021;(July).

27. Waites T. Children and young people’s consultation: Access to health services by children and young people during the COVID-19 pandemic. 2021.

28. Welsh Government. Creating a play friendly Wales. Cardiff; 2012.

29. Moore SA, Faulkner G, Rhodes RE, Brussoni M, Chulak-Bozzer T, Ferguson LJ, et al. Impact of the COVID-19 virus outbreak on movement and play behaviours of Canadian children and youth: A national survey. Int J Behav Nutr Phys Act. 2020;17(1):1–11.

30. Marchant E, Todd C, James M, Crick T, Dwyer R, Kingdom U, et al. Primary school staff reflections on school closures due to COVID-19 and recommendations for the futurel1: a national qualitative survey. 2020.

31. Shum A, Skripkauskaite S, Pearcey S, Walte P, Creswell C. Report 09: Update on children’s & parents/carers’ mental health; Changes in parents/carers’ ability to balance childcare and work: March 2020 to February 2021. 2021;(February):1–22.

32. Alma Economics, Public Health Wales. Children and young people’s mental well-being during the COVID-19 pandemic. London; 2021.

33. James M, Marchant E, Defeyter MA, Woodside J V., Brophy S. Impact of School Closures on the Health and Well-Being of Primary School Children in Wales UK; A Routine Data Linkage Study Using the HAPPEN Survey (2018-2020). SSRN Electron J. 2021;1–19.

34. Proto E, Quintana-Domeque C. COVID-19 and mental health deterioration by ethnicity and gender in the UK. PLoS One [Internet]. 2021;16(1 January):1–16. Available from: http://dx.doi.org/10.1371/journal.pone.0244419

35. Patalay P, Fitzsimons E. Correlates of Mental Illness and Wellbeing in Children: Are They the Same? Results From the UK Millennium Cohort Study. J Am Acad Child Adolesc Psychiatry [Internet]. 2016;55(9):771–83. Available from: http://dx.doi.org/10.1016/j.jaac.2016.05.019

36. Children’s Commissioner for Wales. Coronavirus and Me. 2020;1–68. Available from: https://www.mind.org.uk/information-support/legal-rights/coronavirus-and-your-rights/coronavirus-and-sectioning/

37. Mamot M, Allen J, Goldblatt P, Herd E, Morrison J. Build Back Fairer: The COVID-19 Marmot Review. The Pandemic, Socioeconomic and Health Inequalities in England. London Inst Heal Equity. 2020;

