## Supplementary material for "Factors influencing wellbeing in young people during COVID-19: A survey with 6291 young people in Wales": S1

**Supplementary File 1**

**The HAPPEN Survey**

Consent Form

Before you start please click this link to read the information sheet...

<https://happen-wales.co.uk/childrens-information-sheet/>

1. I have read the child information sheet and understand that if I take part I can change my mind at any time, and this will not be a problem at all. *

*Mark only one oval.*

- Yes
- No

2. I am happy for you to use my questionnaire for research. Only the researchers in the team will know my name and will not tell anyone else my answers *

*Mark only one oval.*

- Yes
- No do not use my questionnaire

3. I am happy for you to look at my school and health records to see how my school is doing (as a group). This is anonymous which means I cannot be identified *

*Mark only one oval.*

- Yes
- No

If you do not wish to take part in the questionnaire please do not continue.

Please click next to start the questionnaire!

ABOUT YOU

4. First Name*

5. Last Name*

6. Home Post Code*

7. What school do you go to?*

8. Do you have any other children living in your house with you (brothers, sisters)?

*Mark only one oval.*

- Yes
- No

9. What year are you in now?*

*Mark only one oval.*

- Year 4
- Year 5
- Year 6

10. Do you have a garden?*

- Yes
- No

11. Gender*

*Mark only one oval.*

- Boy
- Girl
- Prefer not to say

12. Date of Birth

Year*

*Mark only one oval.*

- 2007
- 2008
- 2009
- 2010
- 2011
- 2012

13. Month*

*Mark only one oval.*

- January
- February
- March
- April
- May
- June
- July
- August
- September
- October
- November
- December

14. Day *

*Mark only one oval.*

- 1
- 2
- 3
- 4
- 5
- 6
- 7
- 8
- 9
- 10
- 11
- 12
- 13
- 14
- 15
- 16
- 17
- 18
- 19
- 20
- 21
- 22
- 23
- 24
- 25
- 26
- 27
- 28
- 29
- 30
- 31

YESTERDAY

15. How did you get to school yesterday?*

- On the bus
- On bike
- In the car/taxi
- Walked
- Ran/jogged
- Scooter
- Skateboarded/Rollerbladed

16. What did you have to eat for lunch yesterday?*

- School dinner
- Packed lunch from home
- Nothing

17. What did you do for most of your breaktimes yesterday?*

- Sat around inside or outside
- Ran around
- Stood around
- Walked around

18. How many friends did you play with yesterday?*

- I like to play on my own
- 1-2
- 3-4
- 5 or more

19. Did you have an afternoon break yesterday?*

- Yes
- No

20. How did you get home yesterday?*

- On the bus
- On bike
- In the car/taxi
- Walked
- Ran/jogged
- Scooter
- Skateboarded/Rollerbladed

AFTER SCHOOL

21. How may portions of fruit and vegetables did you eat yesterday?*

- 1
- 2
- 3
- 4
- 5
- 6
- 7
- 8

22. How many times did you brush your teeth yesterday?*

- 0
- 1
- 2
- 3

23. What time did you fall asleep YESTERDAY (to the nearest half hour)?

*Mark only one oval.*

- 6.00pm
- 6.30pm
- 7:00pm
- 7:30pm
- 8:00pm
- 8:30pm
- 9:00pm
- 9:30pm
- 10:00pm
- 10:30pm
- 11:00pm
- 11:30pm
- 12:00am
- 12:30am
- 1:00am
- 1:30am
- 2:00am
- 3:00am
- 3:30am
- 4:00am

24. What time did you wake up TODAY (to the nearest half hour)?

*Mark only one oval.*

- 5:00am
- 5:30am
- 6:00am
- 6:30am
- 7:00am
- 7:30am
- 8:00am
- 8:30am
- 9:00am
- 9.30am
- 10.00am
- 10.30am
- 11.00am
- 11.30am

THE LAST WEEK

25. In the last 7 days, how many days did you do sports or exercise for at least 1 hour in total. This includes doing any activities (this includes any activities or playing sports where your heart beat faster, you breathed faster, and you felt warmer?

*Mark only one oval.*

- 0 days
- 1-2 days
- 3-4 days
- 5-6 days
- 7 days

26. In the last 7 days, how many days did you watch TV/play online games/use the internet etc. for 2 or more hours a day (in total)?

*Mark only one oval.*

- 0 days
- 1-2 day
- 3-4 days
- 5-6 days
- 7 days

27. In the last 7 days, how many days did you feel tired?

*Mark only one oval.*

- 0 days
- 1-2 days
- 3-4 days
- 5-6 days
- 7 days

28. In the last 7 days, how many days did you feel like you could concentrate/pay attention well on your schoolwork?

*Mark only one oval.*

- 0 days
- 1-2 days
- 3-4 days
- 5-6 days
- 7 days
- Don’t do school work

29.  In the last 7 days, how many days did you drink at least one fizzy drink (e.g. coke, fanta, sprite)
 *Mark only one oval.*

- 0 days
- 1-2 days
- 3-4 days
- 5-6 days
- 7 days

30. In the last 7 days, how many days did you eat at least one sugary snack (e.g. chocolate bar, sweets)
 *Mark only one oval.*

- 0 days
- 1-2 days
- 3-4 days
- 5-6 days
- 7 days

31. In the last 7 days, how many days did you eat take away foods (e.g. Chinese takeaway)

*Mark only one oval.*

- 0 days
- 1-2 days
- 3-4 days
- 5-6 days
- 7 days

SPORT AND ACTIVITY

32. These questions are going to ask you how you feel about physical activity (This includes any activity where your heart beats faster, you breathe faster and you feel warmer)

- I want to take part in physical activity
- I feel confident to take part in lots of different physical activities
- I am good at lots of different physical activities
- I understand why taking part in physical activity is good for me

32. How many times do you take part in sports club outside of school a week?

- 0
- 1
- 2
- 3
- 4
- 5
- 6
- 7
- 8
- 9
- 10

33. Can you ride a bike without stabilisers?

- Yes
- No

34. Can you swim 25 metres without a float or armbands? (This is 1 length in a standard swimming pool)

- Yes
- No

YOU AND YOUR FEELINGS

35. Tell us if you agree or disagree with the following:

- I am doing well at school
- I feel part of my school community
- I have lots of choice over things that are important to me
- There are lots of things I’m good at

36. On a scale of 0 to 10 (0 being very unhappy and 10 being very happy), how do you feel about

*Based on the Good Childhood Index by the Children's Society

37. Your Health

*Mark only one oval.*

- 0
- 1
- 2
- 3
- 4
- 5
- 6
- 7
- 8
- 9
- 10

38. Your School

*Mark only one oval.*

- 0
- 1
- 2
- 3
- 4
- 5
- 6
- 7
- 8
- 9
- 10

38. Your Family

*Mark only one oval.*

- 0
- 1
- 2
- 3
- 4
- 5
- 6
- 7
- 8
- 9
- 10

39. Your Friends

*Mark only one oval.*

- 0
- 1
- 2
- 3
- 4
- 5
- 6
- 7
- 8
- 9
- 10

40. Your Appearance

*Mark only one oval.*

- 0
- 1
- 2
- 3
- 4
- 5
- 6
- 7
- 8
- 9
- 10

41. Your Life

*Mark only one oval.*

- 0
- 1
- 2
- 3
- 4
- 5
- 6
- 7
- 8
- 9
- 10

YOU AND YOUR FEELINGS

This part of the survey is going to ask you how you feel. There are no right or wrong answers. You should just pick the answer which is best for you.

Based on the Me and My Feelings Questionnaire (Deighton, Tymms, Vostanis, Belsky, Fonagy, Brown, Martin, Patalay, & Wolpert, 2012)

42. Remember, there are no right or wrong answers, just pick which is right for you.

*Mark only one oval per row.*

I feel lonely

- Never
- Sometimes
- Always

I cry a lot

- Never
- Sometimes
- Always

I am unhappy

- Never
- Sometimes
- Always

I feel nobody likes me

- Never
- Sometimes
- Always

I worry a lot

- Never
- Sometimes
- Always

I have problems sleeping

- Never
- Sometimes
- Always

I wake up in the night

- Never
- Sometimes
- Always

I am shy

- Never
- Sometimes
- Always

I feel scared

- Never
- Sometimes
- Always

I worry when I am at school

- Never
- Sometimes
- Always

I get very angry

- Never
- Sometimes
- Always

I lose my temper

- Never
- Sometimes
- Always

I hit out when I am angry

- Never
- Sometimes
- Always

I do things to hurt people

- Never
- Sometimes
- Always

I am calm

- Never
- Sometimes
- Always

I break things on purpose

- Never
- Sometimes
- Always

YOUR LOCAL AREA

43. On a scale of 0 to 10 (0 being not very safe and 10 being very safe), how safe do you feel playing in your area?

*Mark only one oval.*

- 0
- 1
- 2
- 3
- 4
- 5
- 6
- 7
- 8
- 9
- 10

44. From your house, can you easily walk to school?

*Mark only one oval.*

- Yes
- No

45. From your house, can you easily walk to a park (for example a field, grassy area)?

*Mark only one oval.*

- Yes
- No

45. From your house, can you easily walk to a leisure centre/sports centre?

*Mark only one oval.*

- Yes
- No

46. Can you play in all the places you would like to?

- I can play in all the places I would like to
- I can play in some of the places I would like to
- I can only play in a few places I would like to
- I can hardly play in any of the places I would like to

47. Are you happy with the area that you live in?

- Yes
- No

48. If you could change something to make you and your friends healthier and happier, what would you change... IN SCHOOL?

49. If you could change something to make you and your friends healthier and happier, what would you change... OUT OF SCHOOL?

Don't forget to press submit below!

We have some resources on our website if you would like to learn more or would like to speak to someone... https://happen-wales.co.uk/some-resources-for-you/ (https://happen- wales.co.uk/some-resources-for-you/)

**The COVID & Young People Survey**

Before we get started, are you under 16 or over 16?

1. I am…

- Under 16
- Over 16

Parental Consent

For those aged under 16…

2. I am happy for my child to take part in the COVID & Young People Survey…

- Yes
- No

3. I have read the information sheet about taking part in the study. I have had the opportunity to consider the information, ask questions and have had these answered satisfactorily. I understand that my participation in this research is voluntary and that I am free to withdraw at any time without giving any reason.

I am happy to take part in this online survey...

- Yes
- No

4. Gender

- Female
- Male
- Prefer Not To Say

5. Ethnicity

- Asian
- Black
- Mixed
- White
- Prefer Not To Say

6. Full Home Postcode

7. Date of Birth

8. Are you currently at School/Sixth Form/College or University?

- School
- Sixth Form/College
- University

You and Your School/Sixth Form/College

9. What school/sixth form/college do you go to?

10. What year are you in?

- 7
- 8
- 9
- 10
- 11
- 12
- 13

11. How have you returned after lockdown?

- I am in a small group with a few of my friends
- I am in a smaller group with lots of my friends
- Just smaller class sizes
- I'm not aware there's anything different
- I haven't returned

12. What is your most frequently used method of transport?

- Own car
- Shared car
- School bus
- Public bus
- Train
- Bike
- Walk

13. How long do you travel for per day to get to school/college/sixth form?

- 0 – 10 minutes
- 10 – 20 minutes
- 20 – 30 minutes
- 30 – 40 minutes
- 40 – 60 minutes
- 60+ minutes

14. Do you work?

- No I don’t work
- Part time
- Full time

15. If yes, what is your job?

16. Do any children live in your house?

- Yes
- No

17. How many adults (18+) do you live with at home?

18. Do you have a garden?

- Yes
- No

You and Your University

19. What university do you go to?

20. What type of student are you?

- Undergraduate
- Postgraduate

21. What year are you in?

- First
- Second
- Third
- Fourth
- Fifth

22. Are you a home or international student?

- Home
- International

23. Has your university returned?

- Yes, we are in face to face lectures everyday
- Yes, we are in face to face lectures a few times a week and the rest is online
- Yes, we have smaller face to face seminars/workshops but most is online
- Yes, it is fully online teaching only
- No
- Yes
- Maybe

24. Can you give us any more information about how you feel about your return to university?

25. What type of accommodation do you live in?

- At home with parents/guardians
- Student accommodation on campus
- Student accommodation off campus

26. If you live in student accommodation, is it catered?

- Catered
- Self-catered
- Don’t live in student accommodation

27. Do you live with children?

- Yes
- No

28. How many adults (18+) do you live with?

29. Do you have a garden?

- Yes
- No

30. What is your most frequently used method of transport?

- Own car
- Shared car
- School bus
- Public bus
- Train
- Bike
- Walk

31. How long do you travel for per day to get to university?

- 0 – 10 minutes
- 10 – 20 minutes
- 20 – 30 minutes
- 30 – 40 minutes
- 40 – 60 minutes
- 60+ minutes

32. Do you work?

- No, I don’t work
- Part time
- Full time

33. If yes, what is your job?

In the last week…

34. Have you, or anyone else in your household, had any of the following symptoms in the last 7 days?

- Fever or high temperature
- A cough that has lasted for at least several hours
- Shortness of breath
- Aches and pains, e.g. in back, neck, shoulders or joints
- Blocked nose
- Sore throat
- Feeling unusually tired
- None of these
- I don’t know
- Prefer not to say

35. To the best of your knowledge, do you think you or anyone else in your household have been in direct contact with someone who has COVID19 in the last 7 days?

- Yes
- No
- I don’t know

36. Did you visit any of the following events or locations in the last 7 days?

- School/college/university
- Another family members home
- A friends home
- Pub, bar or café
- Restaurant
- Cinema
- Supermarket or other shop for food or groceries
- Sporting event (as participant), e.g. weekly tennis practice
- Sporting event (as attendee), e.g. a football match
- Indoor location, where over 100 people were present
- Outdoor location, where over 100 people were present
- Religious gathering

37. Have you had a cold in the past week?

- Yes
- No

Yesterday…

38. Did you wear a face mask yesterday?

- Yes
- No

39. If yes, where did you use your face mask?

- Everywhere outside my house
- When walking on the street
- When cycling
- On public transport
- In supermarkets/shops
- In cinema/bar/restaurant
- At home
- At work/school/college/university

40. Do you think wearing a mask is important?

- Yes
- No

41. How much did you spend on your mark? (£)

- 0
- 0-2
- 2-4
- 406
- 6-8
- 8-10
- 10+
- I don’t know

42. How many people including friends/family (who do NOT live in your household) did you meet in person and speak to face-to-face yesterday?

43. How many people including friends/family (who do NOT live in your household) did you meet in person and you had physical contact (e.g. a handshake, hugging, kissing, contact sports).

44. Where did most of this direct contact take place?

- At home
- At school/college/university
- At work

45. Is it easy to remember who you've had direct contact with?

- Yes, I can remember quite accurately how many people I had contact with
- I was guessing a bit, I think I know but I am not sure
- No, it isn't easy to say how many people I had direct contact with

46. If we wanted to have a very accurate answer as to how many people young people have contact with, how would you suggest we do this? Ask people to take a photo of who they met? An app?

47. Would you use this method of keeping track of contacts?

- Yes
- No

You and COVID19…

48. Do you agree or disagree with each of the following statements?

- Coronavirus would be a serious illness for me
- I am likely to catch coronavirus
- If I don’t follow the government’s advice, I might spread coronavirus to someone who is vulnerable
- I am glad schools/colleges/universities have re-opened
- I feel as motivated to learn as I did before lockdown

49. How effective, if at all, do you think these are at slowing the spread of coronavirus?

- Reducing the number of people you meet
- Staying at home for 7 days if you have a mild symptom such as a mild cough
- Staying at home for 7 days if you have more severe symptoms such as a severe cough or a high temperature
- Avoiding crowded places
- Staying at home for 14 days if anyone other than yourself in your household has mild symptom such as a mild cough
- Staying at home for 14 days if anyone other than yourself in your household has severe symptoms such as a cough or a high temperature
- School/college/university closures
- Closing bars, restaurants, cinemas etc.
- Banning the use of public transport

50. On a scale of 1-10, how happy are you with your physical health?

51. Why is this?

52. On a scale of 1-10, how happy are you with your mental health?

53. Why is this?

54. On a scale of 1-10, how happy are you with your life?

55. Why is this?

56. On a scale of 1-10, how happy are you with your family?

57. Why is this?

58. On a scale of 1-10, how happy are you with your friends?

59. Why is this?

60. On a scale of 1-10, how happy are you with your education?

61. Why is this?

62. On a scale of 1-10, how happy are you with your future prospects?

63. Why is this?

64. Do you think anything could have been done differently for young people?

65. Do you think coronavirus has impacted how happy you are with your life?

- Since the coronavirus pandemic, I am happier
- Since the coronavirus pandemic, I am less happy
- Since the coronavirus pandemic, I am the same

Your Wellbeing…

66. Over the last 2 weeks how often have you been bothered by any of the following?

- Feeling nervous, anxious or on edge?
- Not being able to stop or control worrying?
- Worrying too much about different things?
- Trouble relaxing?
- Being so restless that it is hard to sit still?
- Becoming easily annoyed or irritable?
- Feeling afraid as if something awful might happen?

Thank you…

Thank you for taking part in our survey. Please make sure you click next to submit your responses.

We have some resources on our website if you would like to learn more or would like to speak to someone... https://happen-wales.co.uk/some-resources-for-you/

67. Do you have anything else you would like to add?

68. If you are happy for us to contact you if you're successful in our raffle or for a short telephone interview, please leave your e-mail below...
