## Supplementary material for "Factors influencing wellbeing in young people during COVID-19: A survey with 6291 young people in Wales": S2

**Supplementary File 1**

**Variables Used In Analysis**

| The Young People Survey |  |
| --- | --- |
| On a scale of 0 to 10 (0 being very unhappy and 10 being very happy), how do you feel about: *Based on the Good Childhood Index by the Children's Society   - Your health - Your family - Your friends - Your life | Ranked 1 – 10 |
| GAD7 Questionnaire (e.g. Feeling nervous, anxious or on edge?, Not being able to stop or control worrying?) | - Not at all - Several days - More than half the days - Nearly every day |
| The HAPPEN Survey |  |
| Question | Scoring |
| What did you do for most of your breaktimes yesterday? | - Sat around inside or outside - Ran around - Stood around - Walked around |
| How many friends did you play with yesterday? | - I like to play on my own - 1-2 - 3-4 - 5 or more |
| Did you have an afternoon break yesterday? | - Yes - No |
| In the last 7 days, how many days did you do sports or exercise for at least 1 hour in total. This includes doing any activities (this includes any activities or playing sports where your heart beat faster, you breathed faster, and you felt warmer? | - 0 days - 1-2 days - 3-4 days - 5-6 days - 7 days |
| In the last 7 days, how many days did you watch TV/play online games/use the internet etc. for 2 or more hours a day (in total)? | - 0 days - 1-2 days - 3-4 days - 5-6 days - 7 days |
| In the last 7 days, how many days did you feel tired? | - 0 days - 1-2 days - 3-4 days - 5-6 days - 7 days |
| In the last 7 days, how many days did you drink at least one fizzy drink (e.g. coke, fanta, sprite) | - 0 days - 1-2 days - 3-4 days - 5-6 days - 7 days |
| In the last 7 days, how many days did you eat at least one sugary snack (e.g. chocolate bar, sweets) | - 0 days - 1-2 days - 3-4 days - 5-6 days - 7 days |
| I want to take part in physical activity | - Strongly agree - Agree - Disagree - Strongly disagree |
| I feel confident to take part in lots of different physical activities | - Strongly agree - Agree - Disagree - Strongly disagree |
| I am good at lots of different physical activities | - Strongly agree - Agree - Disagree - Strongly disagree |
| I understand why taking part in physical activity is good for me | - Strongly agree - Agree - Disagree - Strongly disagree |
| On a scale of 0 to 10 (0 being very unhappy and 10 being very happy), how do you feel about: *Based on the Good Childhood Index by the Children's Society   - Your health - Your family - Your friends - Your life | - Ranked 1 – 10 |
| Me and My Feelings Questionnaire (e.g., I feel lonely, I cry a lot) | - Never - Sometimes - Always |
