## Supplementary material for "Factors influencing wellbeing in young people during COVID-19: A survey with 6291 young people in Wales": S3

**Supplementary File 2**

**Themes from Thematic Analysis**

| **Theme** | **Quotations** | |
| --- | --- | --- |
| **Primary School** | | |
| **Physical Health** | *“More exercise”*  *“Longer time to play and exercise”*  *“Do more sports”*  *“More PE”*  *“More sports equipment”*  *“Healthy eating”*  *“Eat more fruit and veg”* | |
| **Being with friends** | *“Live closer to my friends”*  *“Being able to play places I want to with my friends”*  *“To play with my friends”*  *“To be with my friends more”* | |
| **Coronavirus concerns** | *“Stop the spread of covid-19 so we can get back to normal”*  *“No covid”*  *“No more covid rules”*  *“For the virus to go away”* | |
| **Secondary School, Sixth Form and University** | | |
| **Mental Health Support** | *“The worsened depression and heightened anxiety caused by coronavirus has almost pushed me to drop out of sixth form”*  *“School for me was never easy but now more than ever that pressure to achieve without the understanding that myself and peers are struggling through an incredibly difficult time and we need to prioritise ourselves and self-care before we even consider the notion of exams in such daunting times”* | |
| **Exam Pressure and Uncertainty** | ***Assessment Concerns*** | *“I am not prepared for GCSE”*  *“Don’t know what is happening with exams etc.”*  *“I never got to sit my a-levels, my grades are much lower than they should be”*  *“Exam situation is very stressful for everyone at the moment ,lack of clarity and answers”*  *“School closures, tension and confusion regarding exams”*  *“At a disadvantage in exams before they even start”*  *“It's extremely difficult to learn with school stopping and starting, my energy levels are much lower, we currently don't know how we are going to be assessed which makes learning difficult as we don't know what we are working towards”*  *“Pressure to well in GCSE's despite missing a lot of school time”* |
|  | ***Feeling Behind and Need for Support*** | *“Struggling to motivate when having to learn at home. Feel I’ve fallen behind”*  *“I am behind in work”*  *“I'm very behind on work and will struggle so much with exams”*  *“The loss of a great deal of education over the lockdown period and huge changes to exams”*  *“Coronavirus has affected my education because I haven't been able to get the support from teachers in the same way I used to”*  *“With less support and more work, many students including myself are struggling to get through the mountainous amount of school work we are assigned”* |
|  | ***Learning from home*** | *“Home learning is hard for me”*  *“We have had to do online learning and that’s impacted our education A LOT”*  *“Online teaching demotivates me from studying because I’m so easily distracted in a home setting”*  *“Unable to have any time/safe space to learn at home means that I cannot produce the quality of work needed to get my education”*  *“My education is now all entirely online, so I've lost access to library services and the interaction of face-to-face teaching”* |
|  | ***Lack of Motivation*** | *“I'm not motivated and I’m just confused”*  *“I can’t focus nor do I feel motivated”*  *“Very hard to stay focused and motivated at all”* |
|  | ***Positives of Distance Learning*** | *“I actually find online learning better, because I can replay the recorded lesson back and make notes in my own time instead of rushing and missing bits of information during class”*  *“I feel like I'm doing more work than I do in school”*  *“I enjoyed working at home”*  *“It has been easier to understand some subjects like Maths because I have had a kind of one to one tuition with my mum*  *My parents made great teachers”* |
| **Future Prospects** | *“Exam cancellation, predicted grades changed, stress of university choices without having to visit them. unsure of my future in education”*  *“I'm currently looking for a specialist college placement and the restrictions have made it much harder to find one at this time”*  *“Delayed me in looking at possible postgraduate routes to take”*  *“I cannot find an Internship”*  *“Constant uncertainty and unknowing of the future, in terms of education it is unknown how A Levels will be assessed fairly across the country to ensure everyone has an equal chance of going to University”* | |
